# Performance of the VISITECT® CD4 Test for Rapid Identification of Advanced HIV Disease at AHF-Supported Sites in Zambia: A Diagnostic Accuracy Analysis in a Zambian Population

**DOI:** 10.1101/2025.01.06.25320078

**Authors:** Eric Mpoyi Mulumba, Mazinga F. Kayembe, Webster C. Chewe, Chitalu Chanda, Nduduzo Dube, Clifford Chituta, Benson M. Hamooya, Freddy T. Kasweka, Martin Matabishi, Muyungila Bob Assani

## Abstract

**Background:** Advanced HIV disease (AHD), defined as a CD4 count below 200 cells/mm³ or the presence of WHO Stage 3 or 4 clinical conditions, poses significant diagnostic challenges in resource-limited settings such as Zambia. Early identification of AHD is essential for improving outcomes for recipients of care (ROCs). The VISITECT® CD4 test, a point-of-care tool, offers potential for addressing these challenges; however, its diagnostic performance in Zambia remains under-evaluated. This study aimed to assess the sensitivity and specificity of the VISITECT® CD4 test for identifying AHD among ROCs at five AIDS Healthcare Foundation (AHF)-supported clinics in Zambia.

**Methodology:** A cross-sectional study was conducted between January 8 to May 31, 2024, at five AHF-supported clinics in Zambia. Participants included individuals who were newly diagnosed with HIV, re-engaging in HIV care, or presenting with unsuppressed HIV viral loads. Baseline clinical and demographic data were collected, and the diagnostic performance of the VISITECT® CD4 test was compared to the flow cytometry CD4 test (the gold standard) using STATA v15 to determine sensitivity, specificity, and other diagnostic accuracy metrics.

**Results:** Among 231 participants, 24.7% (57/231) had AHD based on the flow cytometry CD4 test, whereas 47.6% (110/231) were classified with AHD by the VISITECT® CD4 test. The VISITECT® CD4 test demonstrated a sensitivity of 96.5% and a specificity of 68.4%. The Negative Predictive Value was 98.4%, and the Positive Predictive Value was 50.0%. The overall diagnostic accuracy, as indicated by the area under the curve, was 0.82 (95% CI: 0.78, 0.87). AHD was significantly associated with age according to flow cytometry (p=0.029) but not according to the VISITECT® CD4 test (p=0.277). A significantly higher proportion of individuals with anemia had AHD across both tests (p<0.05).

**Conclusion:** The VISITECT® CD4 test demonstrated strong diagnostic accuracy for detecting AHD, making it a valuable rapid point-of-care tool in resource-limited settings. Its wider implementation could enhance early AHD detection and facilitate timely clinical interventions, warranting further exploration.

## Background

In sub-Saharan Africa (SSA), one of the most pressing challenges in the fight against human immunodeficiency virus (HIV) is the early identification and management of advanced HIV disease (AHD), which continues to drive HIV-related morbidity and mortality [1–4]. The World Health Organization (WHO) characterizes AHD as a CD4 count below 200 cells/mm³ or the presence of a WHO clinical stage 3 or 4 disease in adults and adolescents [2,5–7].

Alarmingly, over 50% of the 1.3 million new HIV infections reported globally in 2022 occurred in SSA where many individuals present late to care, with AHD, putting them at increased risk of opportunistic infections (OIs) such as tuberculosis (TB) and cryptococcal meningitis [7–11]. These OIs significantly contribute to the estimated 10% of people living with HIV who die within the first three months of antiretroviral therapy (ART) initiation, underscoring the urgency of timely AHD diagnosis and management [12–16].

In an effort to reduce AHD-associated mortality, WHO introduced an evidence-based package of care aimed at preventing, diagnosing, and treating common OIs in AHD patients [2,3,16]. A critical obstacle, however, is the availability of simplified, cost-effective point-of-care (POC) diagnostic tools that can identify AHD in asymptomatic individuals—people living with HIV who may not yet display overt symptoms but are at significant risk of life-threatening complications [2]. This gap is particularly pronounced in SSA, where limited access to advanced laboratory services leads to a low diagnostic capacity for CD4 [17,18].

Although HIV management increasingly focuses on viral load testing, the continued need for CD4 testing recommended by WHO highlights the importance of a rapid diagnostic for AHD, especially in settings where conventional CD4 testing is unavailable [16,19]. One innovation is the VISITECT® CD4 test, a lateral flow assay that provides a rapid and reliable method of identifying people living with HIV with CD4 counts below 200 cells/mm³ [20,21]. Despite its potential, data on the test’s sensitivity and specificity in resource-constrained settings, such as Zambia, remain limited. Therefore, this study aimed to evaluate the diagnostic accuracy of the VISITECT® CD4 test in identifying AHD among people with HIV at five AIDS Healthcare Foundation (AHF)- supported sites in Zambia.

## Methods

### Study design and sites

A cross-sectional study was conducted from January 8 to May 31, 2024, at five high-volume ART facilities in Zambia. These facilities—Chifundo and Lusungu AHF ART Clinics in Lusaka District, Kapiri Mposhi Urban Clinic in Kapiri Mposhi District, New Masala Clinic in Ndola District, and Shampande Clinic in Choma District—were purposively selected based on their ability to perform flow cytometry CD4 testing and their large number of ROCs.

### Study population

The study included individuals newly diagnosed with HIV, confirmed by two separate HIV rapid antibody tests in accordance with Zambia’s national HIV guidelines [22], ROCs re-engaging in HIV care after a disengagement period of more than six months, and those who had been on ART for over six months but remained unsuppressed with an HIV viral load of more than or equal to 1,000 copies/ml. For participants under 17 years of age, assent was obtained from an accompanying adult. Exclusion criteria included ROCs who had been on ART for over six months but had a documented viral load below 1,000 copies/ml or no documented viral load. Individuals who declined to provide written informed consent or assent were also excluded.

### Sample size

The sample size of 231 participants was determined by the quantity of VISITECT® CD4 test kits available at each of the five selected facilities during the study period.

### Data Collection

Data were collected using a data abstraction form administered by trained research assistants at each research site. The information gathered included socio-demographic and clinical characteristics such as age, sex, date of HIV diagnosis, date of ART initiation, ART regimen, WHO HIV clinical stage and urine LF-LAM test result. In addition, 8 mL of venous blood were collected from each participant into an EDTA tube. Within 24 hours of collection, several tests were performed on the same specimen, including the CRAG test, RPR test, hemoglobin level measurement, HBsAg test, VISITECT® CD4 test, and flow cytometry CD4 test. The CD4 cell count was measured concurrently using both the VISITECT® CD4 test and flow cytometry. Specifically, the PIMA and BD FacsPresto machines were used to determine the absolute CD4 count via flow cytometry, while the VISITECT® CD4 test was performed according to the specifications and standard operating procedures provided by Omega Diagnostics [23].

### Study definitions

#### Advanced HIV Disease (AHD)

AHD was defined for this study as either a laboratory-confirmed CD4 count below 200 cells/μL or a clinical classification of WHO stage 3 or 4 in individuals living with HIV[2].

#### Antiretroviral Therapy (ART) Status

ART status was categorized into two groups: ART-naïve and ART-experienced. ART-naïve referred to individuals who had never initiated antiretroviral therapy, while ART-experienced included those who had previously been on or were currently on ART treatment.

#### Anaemia

Anaemia was identified as a hemoglobin level of less than 13.0 g/dL for males and less than 12.0 g/dL for females [24].

#### HIV Viral Load

HIV viral load represented the concentration of HIV RNA (viral particles) in the bloodstream, measured in copies per milliliter (copies/mL). Suppressed viral load was defined as viral load less than 200 copies/mL. Low-level viremia (LLV) corresponded to a viral load ranging between 200–999 copies/mL, and unsuppressed viral load referred to levels equal to or exceeding 1,000 copies/mL [2,25].

### Data Processing and analysis

The collected data was initially entered into Microsoft Excel and subsequently exported to Stata version 15 for analysis. For categorical variables, descriptive statistics such as frequency and proportions were calculated to assess data distribution. The Chi-square or Fisher’s exact test was applied to examine associations between AHD and socio-demographic and clinical characteristics. The sensitivity, specificity, and predictive values of the VISITECT® CD4 test were evaluated against the reference methods (flow cytometry CD4 test). The diagnostic accuracy of the VISITECT® CD4 test was assessed using the area under the curve (AUC). Statistical significance was set at p<0.05.

### Ethical considerations

The study protocol received ethical approval from the Tropical Diseases Research Centre’s (TDRC) Ethics Review Committee (TRC/C4/12/2023) and the National Health Research Authority (NHREB006/29/12/2023). Authorization to conduct the study was also obtained from the respective Provincial and District Health Offices, as well as the facility in-charges at each study site. Written informed consent was secured from all participants, and for participants under 18 years of age, written informed consent was obtained from their parent or guardian. Data were collected without any personal identifiers, safeguarded to maintain confidentiality, and made accessible only to the principal and co-principal investigators as appropriate.

## Results

### Characteristics of Study Participants

A total of 231 participants were included, with 24.7% (57/231) having AHD according to the Flow Cytometry CD4 test and 47.6% (110/231) classified as having AHD by the VISITECT® CD4 test. Participants aged 35–45 years constituted the majority of AHD diagnoses, and a significant association between age and AHD was observed with Flow Cytometry (p=0.029), though not with the VISITECT® CD4 test (p=0.277). Most individuals with AHD were ART-naïve, demonstrating a significant association in the VISITECT® CD4 test (p=0.011) but not in Flow Cytometry (p=0.285).

WHO HIV clinical staging was significantly associated with AHD when assessed by Flow Cytometry (p<0.001). The proportion of participants in WHO stages 1, 3, and 4 diagnosed with AHD was 68%, 3.5%, and 5.3%, respectively, using the VISITECT® CD4 test, compared with 79.1%, 4.6%, and 2.7% via Flow Cytometry. Among individuals diagnosed with AHD by Flow Cytometry, 57.9% (33/57) were anemic, compared to 25.4% (44/174) of those without AHD (p<0.001). Similarly, 41.8% (46/110) of participants identified with AHD by the VISITECT® CD4 test were anemic, compared to 25.8% (31/121) of those without AHD (p=0.010).

Notably, AHD was also present among ROCs with suppressed viral loads (<1,000 copies/mL). Specifically, 8.3% (2/24) of such participants were diagnosed using the Flow Cytometry CD4 test, while 14.3% (6/42) were diagnosed using the VISITECT® CD4 test (see Table 1).

**Table 1.**
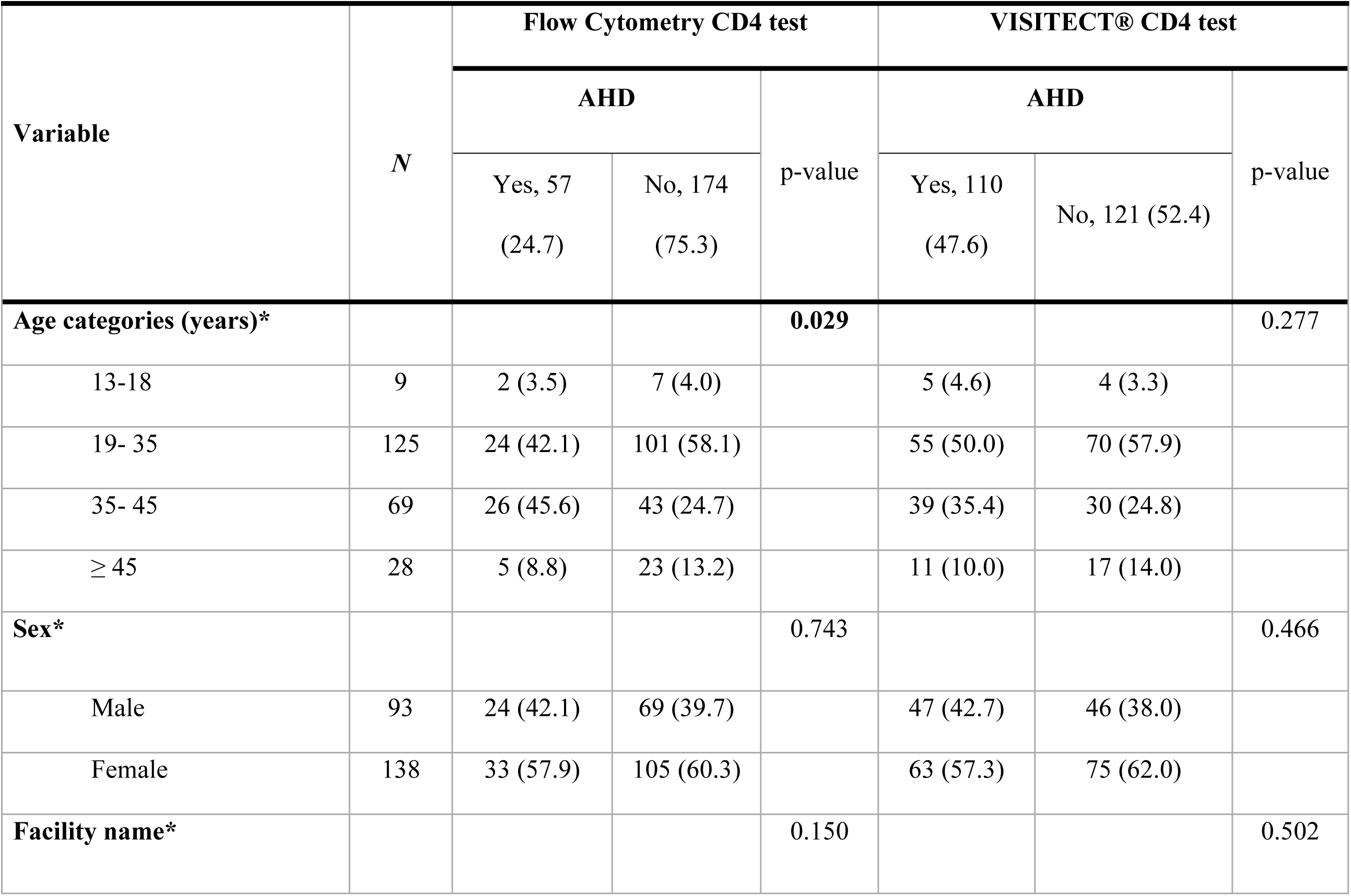
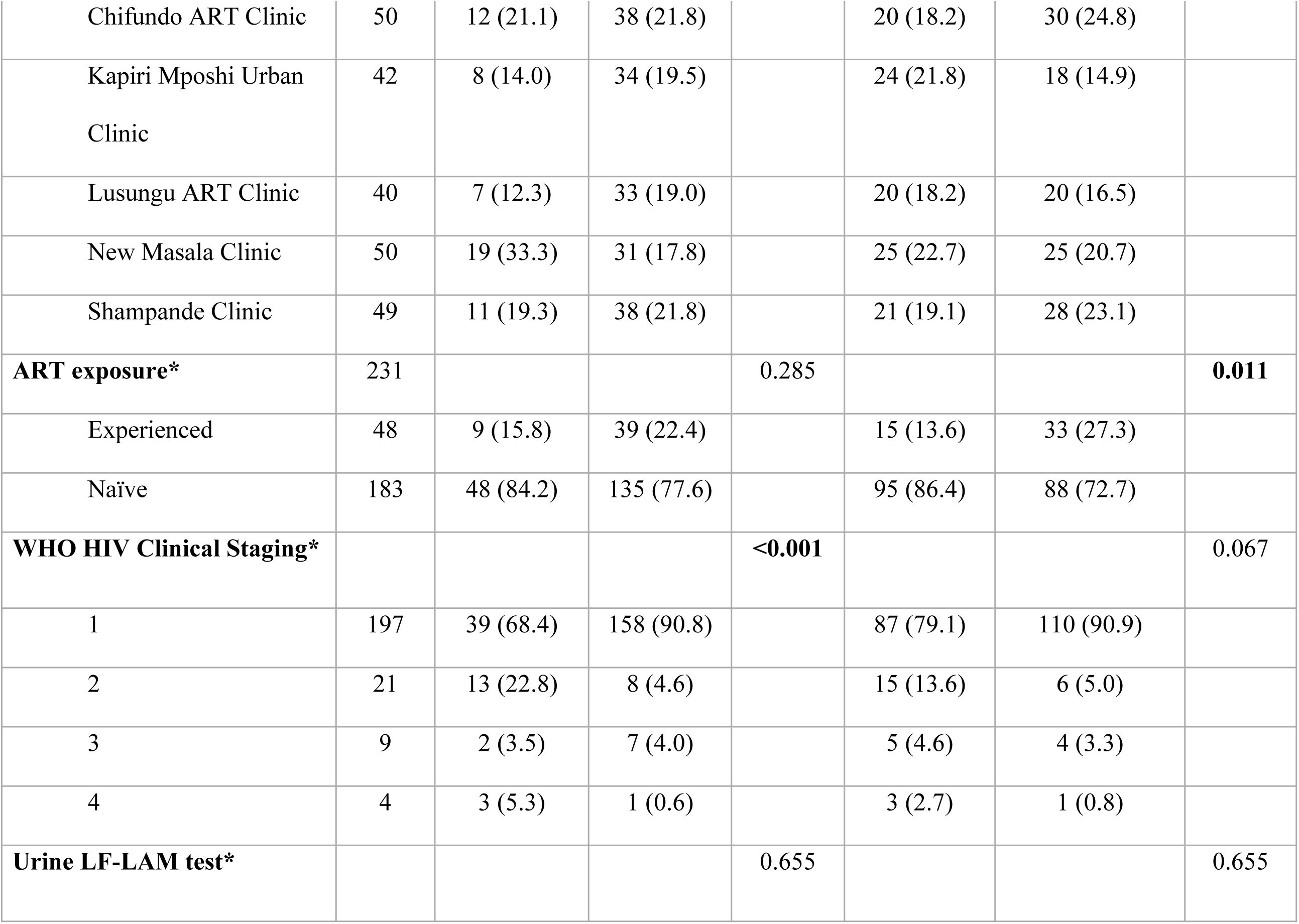
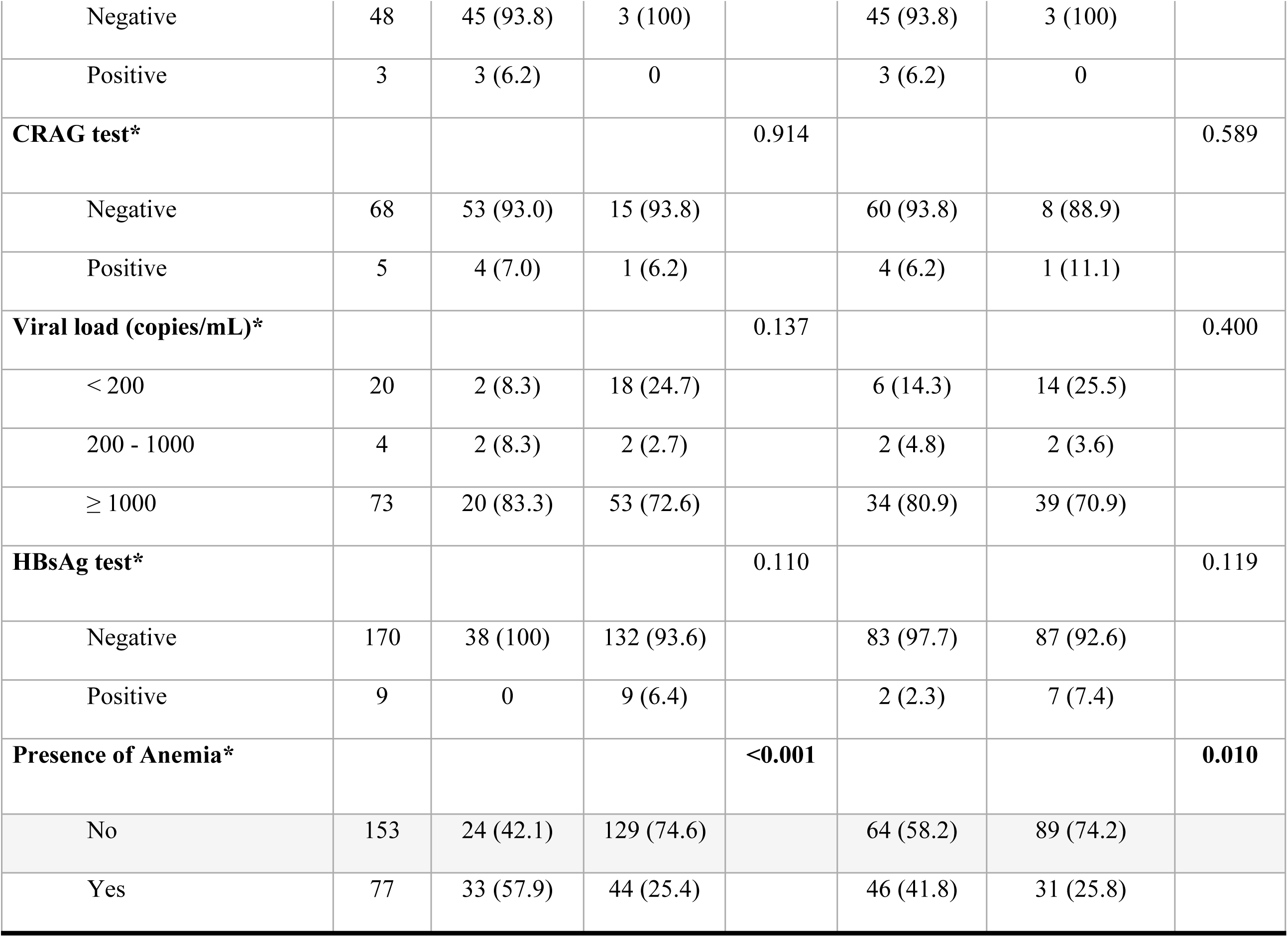
Characteristics of Study Participants and Univariate Comparison by Advanced HIV Disease Status.

### Sensitivity, Specificity, and Diagnostic accuracy of the VISITECT® CD4 test in identifying advanced HIV disease

Table 2 outlines the diagnostic accuracy of the VISITECT® CD4 test compared with the gold standard Flow Cytometry CD4 test in identifying AHD. The VISITECT® CD4 test demonstrated a sensitivity of 96.5% (95% CI: 94.1–98.9) and a specificity of 68.4% (95% CI: 62.4–74.4). The positive predictive value (PPV) was 50.0% (95% CI: 43.6–56.5), while the negative predictive value (NPV) was high at 98.4% (95% CI: 96.7–100). The overall diagnostic accuracy, as measured by the area under the curve (AUC), was 0.82 (95% CI: 0.78–0.87).

**Table 2.**
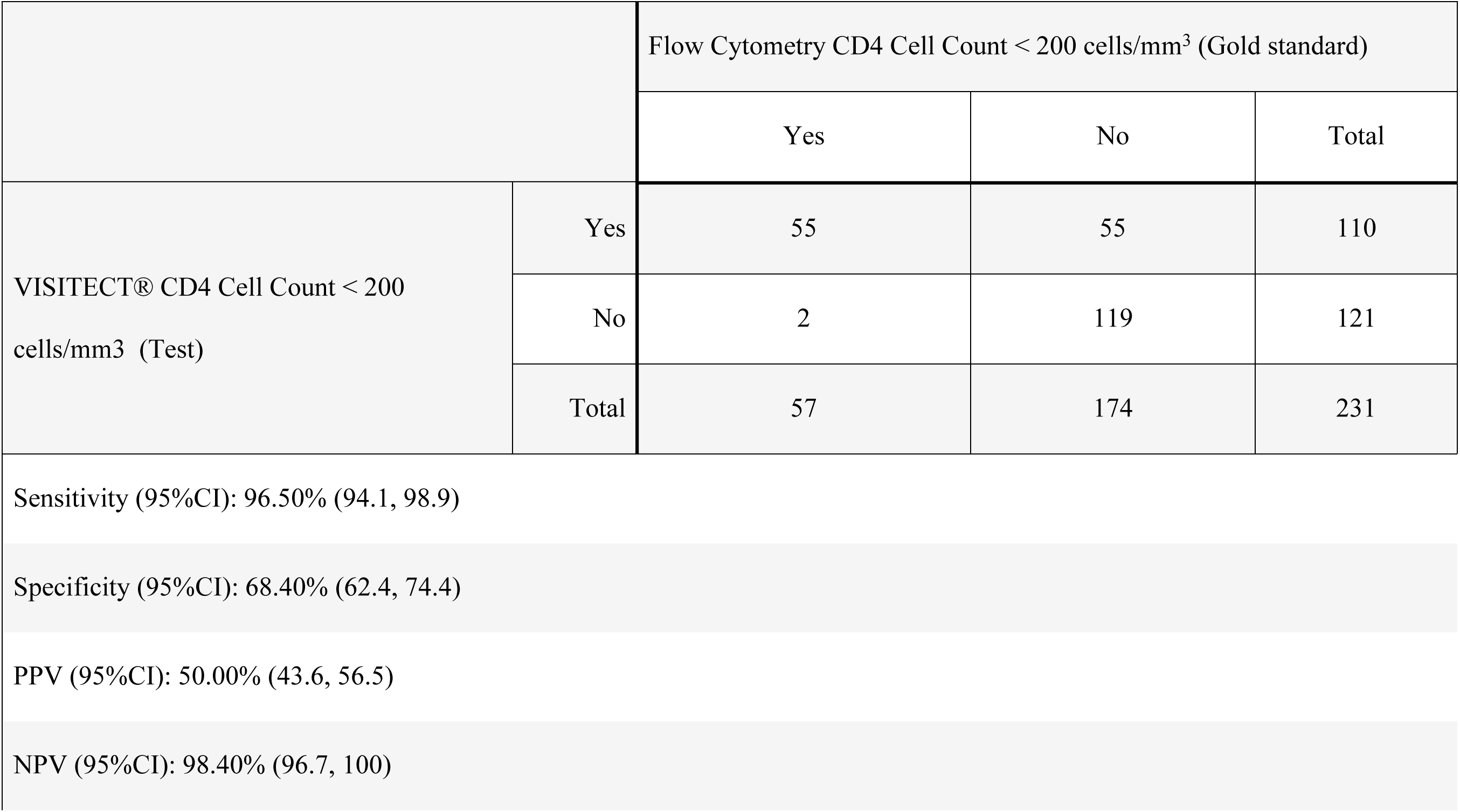
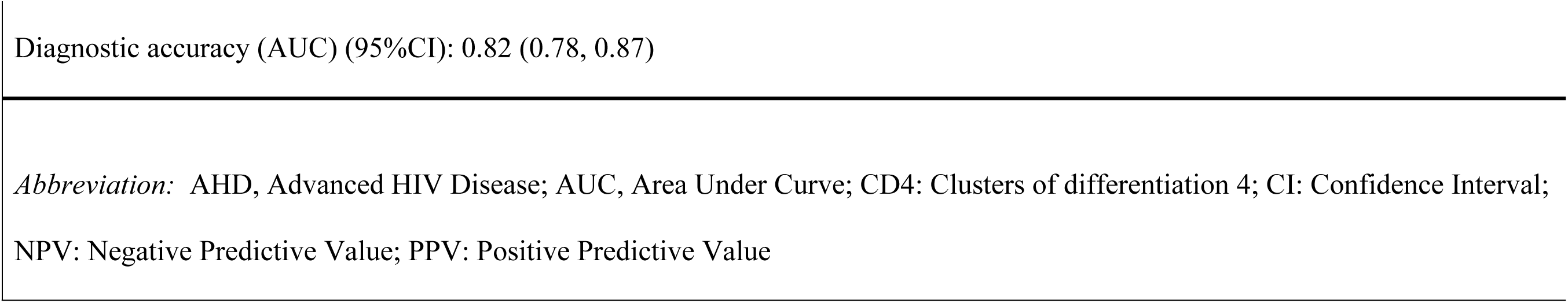
Sensitivity, Specificity, and Diagnostic Accuracy of the VISITECT® CD4 Test Compared to Flow Cytometry for Identifying Advanced HIV Disease.

|  |  | Flow Cytometry CD4 Cell Count < 200 cells/mm<sup>3</sup> (Gold standard) |  |  |
| --- | --- | --- | --- | --- |
|  |  | Yes | No | Total |
| VISITECT® CD4 Cell Count < 200 cells/mm<sup>3</sup> (Test) | Yes | 55 | 55 | 110 |
|  | No | 2 | 119 | 121 |
|  | Total | 57 | 174 | 231 |
| Sensitivity (95%CI): 96.50% (94.1, 98.9) |  |  |  |  |
| Specificity (95%CI): 68.40% (62.4, 74.4) |  |  |  |  |
| PPV (95%CI): 50.00% (43.6, 56.5) |  |  |  |  |
| NPV (95%CI): 98.40% (96.7, 100) |  |  |  |  |

*Abbreviation:* AHD, Advanced HIV Disease; AUC, Area Under Curve; CD4: Clusters of differentiation 4; CI: Confidence Interval; NPV: Negative Predictive Value; PPV: Positive Predictive Value

## Discussion

The study found the sensitivity of the VISITECT® CD4 test to be 96.5% when compared to the Flow Cytometry CD4 test. This is consistent with studies from Zimbabwe and the Democratic Republic of Congo, where similar lateral flow-based CD4 tests demonstrated sensitivities exceeding 95% [20,21,26,27]. The robust performance of the VISITECT® CD4 tes as a POC tool in resource-limited settings provides an opportunity for timely screening of AHD, helping to prevent diagnostic delays that could lead to poor clinical outcomes [21].

However, the specificity of the VISITECT® CD4 test was moderate at 68.4%, indicating a risk of false-positive AHD diagnoses. This may result in unnecessary interventions and increased resource use. A potential explanation for the lower specificity includes over-detection of AHD in PLHIV with borderline CD4 counts and challenges related to suboptimal testing conditions in resource-limited environments. These findings are consistent with reports from Botswana, emphasizing the need for confirmatory testing to mitigate misdiagnoses and reduce unnecessary interventions [20,21,28,29].

The high NPV of 98.4% observed in this study further supports the reliability of the VISITECT® CD4 test in ruling out AHD. This aligns with findings from a multicenter evaluation that reported an NPV of 94.6% highlighting the test’s utility in confidently excluding AHD [30]. By accurately identifying individuals who do not have AHD, the test enables clinicians to allocate limited resources more effectively and focus on those who would benefit most from targeted interventions.

In contrast, the PPV of the VISITECT® CD4 test was relatively low at 50.0%, indicating that half of the individuals identified as having AHD by this test may not truly have the condition. This finding aligns with a laboratory-based validation study in Eswatini, which reported a similar PPV of 48.7% [21,28,29]. The low PPV may reflect the lower prevalence of AHD in the study population and the intrinsic characteristics of lateral flow assays. Targeting populations with a higher pre-test probability of AHD, such as those with unsuppressed viral loads or clinical symptoms, could enhance the test’s predictive value.

The findings underscore the need for a balanced approach to implementing the VISITECT® CD4 test in clinical practice. Its strengths in sensitivity and NPV make it a valuable screening tool for excluding AHD, especially in settings with limited access to flow cytometry. Its rapid turnaround time and simplicity further position it as a practical solution for high-volume clinics, where timely AHD identification is critical.

To maximize its impact, the VISITECT® CD4 test, as a POC tool, should be integrated into a broader diagnostic algorithm, complemented by confirmatory methods for positive cases. Future research should focus on refining the test to improve specificity, assessing its clinical utility across diverse populations, and exploring its effects on operational workflows and patient outcomes. Such studies will provide valuable insights into the test’s cost-effectiveness and scalability, guiding policy decisions and its integration into national HIV programs. By addressing these areas, the VISITECT® CD4 test has the potential to significantly enhance AHD diagnosis and management, reducing HIV-related morbidity and mortality in resource-limited settings. Confirmatory testing remains essential for positive cases to ensure accurate diagnosis and reduce unnecessary interventions.

### Study limitations and strengths

The primary limitation of this study was the relatively small sample size, influenced by the limited availability of VISITECT® CD4 test kits during the study period, which may have reduced the robustness of conclusions, particularly regarding the test’s specificity. Additionally, the study did not explore qualitative aspects such as the impact of the VISITECT® CD4 test on clinician workflows, patient decision-making, or operational challenges like patient waiting times and test integration into clinical settings. Despite these limitations, the study was conducted across diverse high-volume ART facilities reflecting the real-world conditions and included a broad participant population, enhancing its relevance for routine HIV care. Moreover, the use of flow cytometry as a gold standard provided a robust comparison.

## Conclusion

This study demonstrated that the VISITECT® CD4 test is an effective POC diagnostic tool for screening AHD in resource-limited settings, particularly for reliably ruling out AHD in people living with HIV. However, its moderate specificity (68.4%) and lower positive predictive value (50.0%) indicate a risk of overestimating AHD cases, underscoring the need for cautious interpretation of positive results. To enhance diagnostic accuracy, integrating the VISITECT® CD4 test into a comprehensive diagnostic algorithm for AHD is recommended. Future research should prioritize improving the test’s specificity and evaluating its clinical utility in diverse settings, focusing on both quantitative outcomes and qualitative aspects of care delivery.

## Data Availability

No restriction

## Acknowledgments

Authors thank the research teams and study participants from Chifundo and Lusungu AHF ART Clinic, Lusaka, Kapiri Mposhi Urban Clinic, Kapiri Mposhi District, New Masala Clinic, Ndola District and Shampande Clinic, Choma for their valuable time and dedication to ensure this study is a success. Further gratitude goes to the provincial and district health office teams in respective provinces and districts for allowing this research to proceed without any hindrance.

## Author contributions

E.M.M.: Conceptualization, funding acquisition, methodology, supervision, formal analysis, writing of original draft, reviewing and editing draft, finalization of manuscript. M.F.K. and W.C.C.: Conceptualization, methodology, supervision, formal analysis, writing of original draft, reviewing and editing draft, finalization of manuscript. C.C.: Conceptualization, methodology, formal analysis, writing of original draft, reviewing and editing draft, finalization of manuscript. M.M.: Funding acquisition, writing of original draft, reviewing and editing draft, finalization of manuscript. N.D.: Conceptualization, funding acquisition, methodology, formal analysis, writing of original draft, reviewing and editing draft, finalization of manuscript. B.M.H.: Formal analysis, writing of original draft, reviewing and editing draft, finalization of manuscript. F.T.K. and C.C.: Conceptualization, formal analysis, writing of original draft, reviewing and editing draft, finalization of manuscript. M.B.A.: Reviewing and editing draft, finalization of manuscript.

## Data Sharing

The study protocol and statistical code are available from the corresponding author upon request. The data for this study is available from AIDS Healthcare Foundation Zambia.

## Disclaimer

The findings and conclusions in this report are those of the authors and do not represent the official position of the AIDS Healthcare Foundation

